# Evaluation of an artificial intelligence model for opportunistic calculation of Agatston score on non-gated computed tomography of the chest

**DOI:** 10.1101/2024.11.20.24317666

**Authors:** Suzannah E McKinney, Sarah F Mercaldo, John K Chin, Ankita Ghatak, Madeleine A Halle, Sandeep S Hedgire, Nandini M Meyersohn, Brian Ghoshhajra, Keith J Dreyer, Mannudeep Kalra, Bernardo C Bizzo, James M Hillis

**Affiliations:** Mass General Brigham AI, Boston, MA, USA; Department of Radiology, Massachusetts General Hospital, Boston, MA, USA; Harvard Medical School, Boston, MA, USA; Department of Neurology, Massachusetts General Hospital, Boston, MA, USA

**Author notes:** Corresponding author: Suzannah E McKinney, Mass General Brigham AI, 399 Revolution Dr, MA 02145. These authors contributed equally to the work and share last authorship (Bernardo C Bizzo, James M Hillis). Author.

## Abstract

**Importance:** The Agatston score is a measure of cardiovascular disease traditionally calculated on cardiac gated computed tomography (CT) of the chest. Cardiac gated CT is resource-intensive, can be hard to access, and involves extra radiation exposure. Artificial intelligence (AI) can be used to opportunistically calculate Agatston score on non-gated CTs performed for other indications.

**Objective:** This study compared the accuracy of an AI model (Riverain Technologies ClearRead CT CAC) at calculating Agatston scores on non-gated CTs to both consensus radiologist interpretations on the same CTs and Agatston scores from paired cardiac gated CTs.

**Design:** A retrospective standalone performance assessment was conducted on a dataset of non-contrast CT chest cases acquired between January 2022 and December 2023.

**Setting:** The study was conducted at five hospitals in the United States.

**Participants:** The cohort included non-gated CTs from 491 patients. It was enriched to ensure a representation of disease severity by selecting approximately two-thirds of patients using the originally reported Agatston score on a paired cardiac gated CT within the study timeframe.

**Main Outcome(s) and Measure(s):** The study compared the agreement of Agatston categories (0, 1-99, 100-399 and ≥400) between the AI model and ground truth radiologists or original radiology reports using the quadratic weighted Kappa coefficient.

**Exposure(s):** Each non-gated CT case was interpreted independently by three radiologists to establish consensus interpretations. Each CT was then interpreted by the AI model. The Agatston scores for paired cardiac gated CTs were obtained from original radiology reports.

**Results:** The agreement between the AI model and ground truth radiologists was 0.959 (95% CI: 0.943-0.975). This result was broadly consistent across sex, age group, race, ethnicity and CT scanner manufacturer subgroups. The agreement between the AI model and paired cardiac gated CT was 0.906 (95% CI: 0.882-0.927).

**Conclusions and Relevance:** The assessed AI model accurately calculated Agatston scores on non-gated CTs and produced similar scores to paired cardiac gated CTs. Its use could broaden screening for atherosclerotic cardiovascular disease, enabling opportunistic screening on CTs captured for other indications.

## Introduction

The prevalence of cardiovascular disease (CVD) in adults in the United States is 48.6% (127.9 million people).^1^ Heart disease and stroke claim more lives each year than cancer and chronic lower respiratory disease combined. Moreover, CVD has huge economic impacts with the annual direct and indirect costs estimated to be $407.3 billion in the United States in 2018 to 2019. Morbidity and mortality can be significantly reduced through primary and secondary prevention strategies, guided by individualized risk assessments. This approach is particularly important when patients remain asymptomatic,^2^ although a challenge for many asymptomatic patients is that they may not have undergone as extensive diagnostic work-up to establish their risk assessment.

Computed tomography (CT) coronary artery calcium (CAC) scoring provides an effective, noninvasive method for predicting CVD. It involves the measurement of Agatston score, which is calculated by multiplying the area of calcifications by a factor derived from their highest attenuation value and then obtaining the sum of these products.^3^An Agatston score can then be categorized into four groups for disease stratification. This technique has demonstrated its ability to reliably assess risk across various populations and provides particular value for prognostication of patients classified as intermediate risk by traditional risk models.^4,5^ One research group followed a total of 42,224 patients of diverse races and ethnicities for a median of 11.7 years; they found that CAC severity based on Agatston category (0, 1-99, 100-399 and ≥400) correlated with risk of all-cause and CVD mortality in all studied race/ethnicity groups. ^6^Further, CAC severity was correlated with all cause and CVD mortality in groups who may be poorly represented in patient cohorts informing the standard risk prediction models otherwise recommended by professional bodies.^6^ Other large cohort studies have further established Agatston scoring as a valuable tool to assess future CVD risk across different age and ethnicity subgroups.^7–9^

A key challenge with CT CAC measurement is that it has traditionally involved gating the acquisition of the CT with cardiac rhythm to reduce motion artifact. This approach demands specialized training of radiology technicians and requires patients to undergo a CT for this limited purpose with the associated radiation exposure.^10^ However, there is the opportunity to use routine, non-gated chest CTs that are obtained for non-cardiac indications, which show CAC but whose presence is not routinely reported.^11^ A 2024 meta-analysis of 108 studies concluded that identification of incidental CAC on non-gated thoracic CT is a useful technique given the occurrence of CAC on non-gated scans was both related to the presence of traditional cardiovascular risk factors (except for smoking status and body mass index) and was predictive of the development of subsequent cardiovascular events and all-cause mortality.^12^

A further challenge is the time required by radiology technicians or radiologists to create manual segmentations of calcified regions to calculate an Agatston score. Artificial Intelligence (AI) has been used opportunistically in medical imaging elsewhere^13^ and the use of AI to automate manual segmentation has been proposed to enable opportunistic screening on non-gated chest CT. This study assessed the performance of an AI device (Riverain Technologies ClearRead | CT CAC), which was designed to estimate the Agatston score by accurately segmenting CAC on non-contrast, non-gated chest CT.

This study aimed to assess the performance of this AI device by comparing it to the Agatston score and Agatston category obtained from ground truth radiologists on the same non-gated chest CT. For a subset of CTs, it also compared the device output to the Agatston score obtained from a paired gated CT. This combined approach allowed evaluation of the device compared to both a manual approach on the same non-gated CT and the clinical gold standard on the gated CT.

## Methods

### Study design

This retrospective standalone model performance study was conducted using radiology cases from five hospitals within the Mass General Brigham (MGB) network between January 2022 and December 2023. It was approved by the MGB Institutional Review Board with waiver of informed consent. It was conducted in accordance with relevant guidelines and regulations including the Health Insurance Portability and Accountability Act (HIPAA). This report followed the Standards for Reporting Diagnostic Accuracy (STARD 2015) reporting guideline.The data from this study include protected health information; some data may be available for research purposes from the corresponding author upon reasonable request.

### Case selection

The study cohort was selected using two distinct methods. The first method aimed to ensure representation across the four Agatston categories (0, 1-99, 100-399 and ≥400) by taking equal numbers of cases in each category based on original radiology reports. The second method aimed to represent prevalence of CAC amongst the general population by taking cases regardless of the Agatston category. For both methods, the cohort considered non-gated CT chest cases performed in any setting (i.e., inpatient or outpatient); there were no limitations on the original CT chest clinical indication. These cases were obtained from patients at least 30 years of age. Repeat cases from the same patient were removed with only the earliest one maintained.

For the first method, a list of paired cases was created, where patients had received both a gated CT chest and non-gated CT chest within the study time period (average duration (± standard deviation) between the two studies of 219.0 ± 175.1 days). The reason for this pairing was that the Agatston score is typically only reported as part of a gated chest CT. The paired approach therefore enabled enrichment of the dataset to ensure the presence of cases across the spectrum of CAC severity; albeit the non-gated chest CT cases were ultimately used in the study. These cases were ordered prospectively by date and then selected consecutively for each of the four Agatston score categories, in a balanced manner across the hospitals, until there were 83 cases in each category (i.e., 332 cases total).

For the second method, a list of non-gated chest CT cases was created for patients that did not have a paired gated chest CT case (i.e., could not be selected using the first method). These cases were ordered prospectively by date and then selected consecutively, irrespective of Agatston score, until there were 33-34 cases at each of the five hospitals (i.e., 168 cases total).

### Image quality review and series selection

All cases were deidentified and then underwent an image quality review by an American Board of Radiology (ABR)-certified radiologist. Cases were excluded if they contained evidence of prior cardiac surgery, presence of intravenous or oral contrast, presence of para-cardiac metal artifact, presence of significant motion artifact that precluded assessment of CAC or presence of significantly altered thoracic anatomy.

At the same time as the image quality review, the radiologist selected the series to be given to the model. To facilitate a range of series slice thicknesses, the radiologist selected up to two soft or standard kernel axial series as available: one that was >0.5mm and ≤1.5mm, and one that was >1.5mm and ≤3mm. When a case had both series available and they both passed the DICOM metadata review as described below, a single series was subsequently selected using randomization that ensured an even balance between these two slice thickness ranges. The ground truth radiologists and the AI model only received this single selected series. The image quality review and series selection process was performed using the FDA-cleared eUnity image visualization software (Version 6 or higher) and an internal web-based annotation system that utilized the REDCap electronic data capture tools hosted at MGB.^14,15^

### DICOM metadata review

A DICOM metadata review was performed after series selection to ensure that the slice thickness was within the appropriate range (>0.5mm and ≤1.5mm, >1.5 and ≤3mm), the slice thickness was consistent (tolerance of 0.01mm), the slice interval was consistent (tolerance of 0.01mm), the slice thickness was equal to or greater than the slice interval (i.e., there were no ‘gaps’ between slices), and the pixel spacing was <1mm.

### Ground truth interpretations

The ground truth interpretations were performed by three ABR-certified radiologists. They independently segmented the regions of CAC using the TeraRecon iNtuition software (version 4.7.0.22-111). As part of this process, they identified the region of CAC as being in the right coronary artery, left anterior descending artery, left circumflex artery and left main coronary artery. The software then calculated the volume of these regions and multiplied them by the Agatston factor to achieve individual Agatston scores for each vessel as well as an overall Agatston score.^3^ The continuous Agatston scores were also categorized using established categories: 0, 1-99, 100-399 and ≥400.^16^ We note that Agatston score is traditionally calculated based on the area on 2.5mm or 3mm slice thickness series from cardiac gated CT; the volume calculated here was scaled to match 3mm series. In addition, for ease of explanation, we have referred to “Agatston score” for any measurements of CAC that incorporate a volume or area multiplied by the Agatston factor while recognizing that its strictest definition only applies to the overall score obtained on cardiac gated CT.

The subsequent analyses used the average of the three ground truth radiologists for the continuous Agatston scores (including overall and for each of the four vessels). The analyses for the overall Agatston category used a consensus category, which was the most frequent category amongst the three ground truth radiologists. All cases had a category with at least two ground truth radiologists (i.e., there were no cases where each radiologist recorded a different category). The segmentation masks from each individual radiologist were also used; they were filtered to only include voxels with ≥130 Hounsfield units that would be used for Agatston calculations.

### Model inference

The evaluated AI model was version 1.1.1.37 of the Riverain Technologies ClearRead | CT CAC device. In brief, it consisted of a U-Net style encoder-decoder architecture that operates across the slices labeling calcified components into one of several categories, including aortic wall and valve calcifications.^17^ The model segments CAC, normalizes for series abnormalities and calculates an Agatston score. It was trained on approximately 2,000 chest CTs with varied protocols and acquisition parameters. The model was installed at MGB for use in this study and received only the relevant series from the Digital Imaging and Communications in Medicine (DICOM)-formatted non-gated CT chest cases. It outputted a continuous score for each vessel (right coronary artery, left anterior descending artery, left circumflex artery and left main coronary artery), an overall continuous Agatston score, and an overall Agatston category. The model also provided an intermediate output of the segmentation masks for any identified areas of CAC.

### Statistical analysis

The statistical analysis was performed in R (version 4.0.2) on the full analysis set. The outputs used for the statistical analysis included the ground truth radiologist interpretations from the non-gated CT scans, the model outputs from the non-gated CT scans, and the previously reported Agatston scores from paired gated CT scans. In addition, demographic and technical parameters were derived from clinical databases or DICOM fields for subgroup analyses. Any missing data were treated as “Unknown” and no data were imputed. All analyses had 95% confidence intervals (CIs) calculated using bootstrapped intervals with 2,000 resamples.

The predefined primary endpoint was the accuracy of the model in calculating the Agatston category as defined by the quadratic weighted Kappa coefficient when compared with the consensus ground truth radiologist Agatston category. The percentage accuracy (i.e., proportion of cases with matching categories) was also calculated. The sample size had been determined based on powering to ensure the lower bound of the 95% CI of the Kappa coefficient was at least 0.85 using preliminary results.

The predefined analyses for comparing the model to the ground truth radiologist interpretations included the accuracy of the overall and component vessel continuous Agatston scores as defined by the Spearman correlation coefficient. There were also predefined subgroup analyses of the overall Agatston categories and scores for sex, age, race, ethnicity, CT scanner manufacturer and radiation dose protocol. Analyses were performed for comparing the paired gated CT overall Agatston categories and scores to both the model outputs and ground truth radiologist interpretations. These analyses continued to use the quadratic weighted Kappa coefficient and percentage accuracy for Agatston categories, and Spearman correlation coefficient for the Agatston scores.

Additional analyses were conducted to compare the agreement of segmentation masks between the model and ground truth radiologist interpretations using Dice scores. This comparison calculated the Dice score between the device and each of the ground truth radiologists (i.e., device versus radiologist 1, device versus radiologist 2, device versus radiologist 3) as well as between each pair of the ground truth radiologists (i.e., radiologist 1 versus radiologist 2, radiologist 2 versus radiologist 3, radiologist 1 versus radiologist 3). The median Dice score for each set of three measurements was used for a case when all three ground truth radiologists provided segmentation; the mean Dice score was otherwise used. The overall average Dice score for the device versus ground truth radiologists and between ground truth radiologist pairs was then calculated together with 95% CIs. The difference between the average Dice scores was also calculated. A non-parametric Wilcoxon rank-sum test tested whether the distributions of Dice scores for model versus ground truth radiologists and between ground truth radiologist pairs were significantly different.

## Results

The cohort for this study included 491 cases and the model successfully performed inference on all cases (Figure 1). This cohort included 272 (55.4%) women and 219 (44.6%) men; the mean (standard deviation) age was 65.6 (10.1) years (Table 1). There were 143 cases in Agatston category 0, 125 in Agatston category 1-99, 84 in Agatston category 100-399 and 139 in Agatston category ≥400.

**Table 1:** Demographic and technical breakdown of CT chest cases.

|  | **Agatston score**<br>**0** | **Agatston score**<br>**1-99** | **Agatston score**<br>**100-399** | **Agatston score**<br>**≥400** | **Overall** |
| --- | --- | --- | --- | --- | --- |
| **Total cases** | 143 | 125 | 84 | 139 | 491 |
| **Sex** |  |  |  |  |  |
| Male | 42 (29.4%) | 50 (40.0%) | 39 (46.4%) | 88 (63.3%) | 219 (44.6%) |
| Female | 101 (70.6%) | 75 (60.0%) | 45 (53.6%) | 51 (36.7%) | 272 (55.4%) |
| **Age** |  |  |  |  |  |
| ≤65 years | 95 (66.4%) | 70 (56.0%) | 40 (47.6%) | 46 (33.1%) | 251 (51.1%) |
| >65 years | 48 (33.6%) | 55 (44.0%) | 44 (52.4%) | 93 (66.9%) | 240 (48.9%) |
| Age, Mean ± SD | 61.8 ± 9.7 | 64.2 ± 8.6 | 66.8 ± 10.0 | 69.9 ± 10.1 | 65.6 ± 10.1 |
| **Race** |  |  |  |  |  |
| Asian | 6 (4.2%) | 3 (2.4%) | 1 (1.2%) | 1 (0.7%) | 11 (2.2%) |
| Black or African American | 4 (2.8%) | 3 (2.4%) | 0 (0.0%) | 0 (0.0%) | 7 (1.4%) |
| Other | 4 (2.8%) | 4 (3.2%) | 2 (2.4%) | 1 (0.7%) | 11 (2.2%) |
| Unavailable | 0 (0.0%) | 0 (0.0%) | 1 (1.2%) | 1 (0.7%) | 2 (0.4%) |
| White | 125 (87.4%) | 113 (90.4%) | 78 (92.9%) | 132 (95.0%) | 448 (91.2%) |
| Declined | 4 (2.8%) | 2 (1.6%) | 2 (2.4%) | 4 (2.9%) | 12 (2.4%) |
| **Ethnicity** |  |  |  |  |  |
| Hispanic | 6 (4.2%) | 2 (1.6%) | 3 (3.6%) | 1 (0.7%) | 12 (2.4%) |
| Not Hispanic | 129 (90.2%) | 116 (92.8%) | 76 (90.5%) | 131 (94.2%) | 452 (92.1%) |
| Unavailable | 5 (3.5%) | 6 (4.8%) | 3 (3.6%) | 6 (4.3%) | 20 (4.1%) |
| Prefer Not to Say/Declined | 3 (2.1%) | 1 (0.8%) | 2 (2.4%) | 1 (0.7%) | 7 (1.4%) |
| **Manufacturer** |  |  |  |  |  |
| GE Medical Systems | 56 (39.2%) | 54 (43.2%) | 46 (54.8%) | 60 (43.2%) | 216 (44.0%) |
| Phillips | 5 (3.5%) | 2 (1.6%) | 5 (6.0%) | 8 (5.8%) | 20 (4.1%) |
| Siemens | 71 (49.7%) | 61 (48.8%) | 27 (32.1%) | 55 (39.6%) | 214 (43.6%) |
| Toshiba | 11 (7.7%) | 8 (6.4%) | 6 (7.1%) | 16 (11.5%) | 41 (8.4%) |
| **Radiation dose protocol** |  |  |  |  |  |
| Low dose | 74 (51.7%) | 69 (55.2%) | 54 (64.3%) | 72 (51.8%) | 269 (54.8%) |
| Routine dose | 69 (48.3%) | 56 (44.8%) | 30 (35.7%) | 67 (48.2%) | 222 (45.2%) |
| **Paired cardiac-gated CT** |  |  |  |  |  |
| No paired gated scan | 37 (25.9%) | 37 (29.6%) | 24 (28.6%) | 62 (44.6%) | 160 (32.6%) |
| Paired gated scan | 106 (74.1%) | 88 (70.4%) | 60 (71.4%) | 77 (55.4%) | 331 (67.4%) |
| Days between gated and non-gated scans | 216.5 ± 184.9 | 226.4 ± 163.7 | 206.4 ± 167.8 | 223.9 ± 181.8 | 219.0 ± 175.1 |

**Figure 1:** Cohort diagram.

The agreement of the overall Agatston category based on quadratic weighted kappa between the model and consensus ground truth interpretations was 0.959 (95% CI: 0.943 to 0.975) with accuracy of 92.3% (95% CI: 89.9% to 94.6%; Figure 2). The correlation of the overall continuous Agatston score between the model and the average ground truth radiologist interpretations as defined by the Spearman correlation coefficient was 0.975 (95% CI: 0.962 to 0.987). The agreement was broadly consistent across sex, age, ethnicity, race, manufacturer and radiation dose protocol subgroups. All subgroups had a kappa coefficient of at least 0.90 except for the “Other” race subgroup, which had a kappa coefficient of 0.855 for only 11 cases (Table S1).

**Figure 2:** Comparison of AI model output compared with ground truth radiologists on non-gated CT for the Agatston category (A) and Agatston score (B).

The correlation of Agatston scores for individual arteries showed a Spearman coefficient of 0.951 (95% CI: 0.925 to 0.971) for the right coronary artery, 0.985 (95% CI: 0.978 to 0.991) for the left anterior descending artery, 0.867 (95% CI: 0.824 to 0.904) for the left circumflex artery and 0.791 (95% CI: 0.736 to 0.839) for the left main coronary artery (Figure 3).

**Figure 3:** Scatterplots comparing AI model output with ground truth radiologists on non-gated CT for the Agatston score for each individual vessel including the right coronary artery (A), left anterior descending artery (B), left circumflex artery (C) and left main coronary artery (D).

The Dice scores were calculated based on 368 segmentations available from the model and 291, 287 and 288 segmentations from the three ground truth radiologists. The average Dice score between the model and ground truth radiologists was 0.892 (95% CI: 0.871 to 0.911) and the average Dice score between ground truth radiologists was 0.910 (95% CI: 0.893 to 0.927). The average difference between Dice scores was -0.008 (95% CI: -0.024 to 0.007), and there was no evidence of a difference between the distributions of Dice scores (p = 0.12).

When considering the agreement of the Agatston categories between the model output on the non-gated CT and the reported values for the paired cardiac gated CT, the kappa coefficient was 0.906 (95% CI: 0.882 to 0.927) with accuracy of 79.2% (95% CI: 74.8% to 83.4%; Figure 4). The Spearman correlation coefficient of the Agatston score was 0.942 (95% CI: 0.920 to 0.957). The comparisons between the ground truth radiologist interpretations on the non-gated CT and the reported values for the paired cardiac gated CT had kappa coefficient of 0.907 (95% CI: 0.883 to 0.930), accuracy of 79.5% (95% CI: 75.2% to 83.6%) and Spearman correlation coefficient of 0.941 (95% CI: 0.920 to 0.957; Figure S1).

**Figure 4:** Comparison of AI model output on non-gated CT with previously reported Agatston categories (A) and scores (B) from paired cardiac gated CT.

## Discussion

This retrospective study assessed the performance of an AI model at quantifying CAC using the Agatston method on non-gated CT. Its accuracy was compared with both ground truth radiologist interpretations of the same non-gated CT and Agatston scores from the original radiology report of paired cardiac gated CTs. The model achieved quadratic weighted kappa coefficients of 0.959 and 0.906 for the respective comparisons for the Agatston categories. It achieved Spearman correlation coefficients of 0.975 and 0.942 for the respective comparisons for the Agatston scores.

These results demonstrate that the model can accurately quantify Agatston scores on non-gated CTs. They show a potential clinical use in screening for incidental coronary artery disease during routine CTs. Importantly, these results also appear to generalize across demographic and technical subgroups, a critical prerequisite for a model to perform consistently across diverse populations.^6^ The economic benefits of cardiac gated CTs versus more invasive measures such as cardiac angiography have previously been shown, as have the benefits of risk stratification and modification compared to untreated cardiovascular disease.^10,18^ It may be possible to realize even greater benefits given the higher number of people undergoing routine CTs compared to cardiac gated CTs. This AI model could therefore truly enable opportunistic screening for atherosclerotic cardiovascular disease.

Validation studies that compare AI model quantification of coronary artery calcium on non-gated CT to ground truth results from cardiac gated CT have been reported. The FDA-cleared AVIEW CAC device by Coreline^19^ was assessed on a cohort of 567 patients for five categories of Agatston scores (it used the categories 0, 1-10, 11-100, 101-400 and >400); it specifically used low-dose non-gated CT.^20^ This model achieved Cohen’s weighted kappa of 0.809 (95% CI: 0.776 to 0.838) with slice thickness 1mm and 0.776 (95% CI: 0.740 to 0.809) with slice thickness 2.5mm. A separate model, which involved team members from Bunkerhill who have subsequently received FDA clearance for the iCAC device^21^, achieved kappa coefficients of 0.802, 0.684, 0.644 and 0.583 on a cohort of 303 cases from four different sites.^22^ The current model compared favorably by achieving a kappa coefficient of 0.906 (95% CI: 0.882 to 0.927) for a similar analysis.

Although these previous works generate valuable proof points about the validity of using AI models to calculate Agatston scores from non-gated CT, and therefore the enablement of opportunistic screening on non-gated CT, our study here goes further to demonstrate the viability of opportunistic screening scoring on non-gated studies in three important ways. Firstly, we account for acquisition parameter differences between gated and non-gated scans. As the 2021 paper points out, because non-gated chest CT are performed for non-cardiac indications, acquisition parameters differ for gated coronary CTs which can affect the accuracy of calcium quantitation. Our study intentionally includes cases from both low-dose and routine-dose protocols, as well as varying slice thicknesses, to minimize the impact of acquisition parameter differences when comparing gated and non-gated scans. Secondly, we show stronger correlation between the AI model Agatston score output and consensus ground truth than previously seen. Thirdly, we used as our primary source of ground truth Agatston scores derived from the manual segmentations of three radiologists on a large non-gated dataset of 491 patients.

Although agreement was high all around, there was higher agreement in our study [0.959 (95% CI: 0.943-0.975)] between both the AI model output and consensus ground truth than AI model and Agatston scores from paired gated studies [0.906 (95% CI: 0.882-0.927)] and between consensus ground truth and Agatston scores from paired gated studies [0.907 (95% CI: 0.883, 0.930)]. Although CAC develops very slowly over time and time periods of <1 year are unlikely to lead to disparate changes between studies, there may be some small impact from the temporal delay between paired studies. Perhaps more importantly, non-gated studies offer a higher spatial resolution than gated due to a thinner typical slice thickness; it is therefore possible that non-gated studies detect calcium that is not seen in gated studies. However, non-gated scans may contain cardiac motion that obfuscates calcium measurement and contain generally more noise, which may quantification of calcium more challenging.

At a coronary vessel level, the model showed better performance for the right coronary artery and left anterior descending artery (Spearman correlation coefficient of 0.951 and 0.985 respectively) than for the left circumflex artery and left main coronary artery (0.867 and 0.791 respectively). A key possible reason is that the left circumflex artery and left main coronary artery are more susceptible to cardiac motion and therefore more challenging to segment. The left circumflex artery (LCX) and left main coronary artery (LM) are particularly affected by respiratory and cardiac motion due to their lateral and anterior positions, with the LCX being more tortuous and susceptible to motion artifacts. Additionally, the LCX’s smaller, more curved vessels experience greater displacement from diaphragmatic motion, while the LM is influenced by the significant movement of the heart’s base during the cardiac cycle. In addition, the delineation of the left circumflex artery and the left main coronary artery may not be as clear, which may cause inconsistencies between the ground truth radiologists and the model in attributing calcium to either of these vessels. This situation may reflect why the Agatston score correlation overall (i.e., based on the sum of the four vessels) remains high despite the lower correlations for these specific vessels.

A key limitation of this study is that it is a retrospective study outside of the clinical workflow. This study therefore establishes the accuracy of the model but does not assess its impact on the clinical workflow and ultimately on clinical outcomes. Key future research questions include whether the model decreases the need for separate gated cardiac CT cases, how consistently its outputs are incorporated into the radiology reports for non-gated cardiac CT cases and whether those outputs lead to changes in preventative strategies for coronary artery disease.

In conclusion, this study demonstrates that this AI model accurately quantifies coronary artery calcium on non-gated CTs, with strong agreement when compared to both ground truth radiologist interpretations and paired gated CT Agatston scores and categories. Its use in routine non-gated CT scans presents a significant possibility for opportunistic screening, enabling earlier detection of coronary artery disease without the need for specialized cardiac imaging. It could therefore broaden access to screening, facilitate timely intervention and ultimately improve cardiovascular health outcomes.

## Supporting information

Figures (Main text)

## Data Availability

Data is not publicly available as it contains Protected Health Information (PHI). We do not have IRB approval for public data-sharing.

## Acknowledgements

The authors thank the broader Mass General Brigham AI and Riverain Technologies teams for their assistance with this project.

## Supplemental Material

Figure S1, Table S1

## Figure Legends

**Table S1:** Subgroup analyses for demographic and technical variables for the agreement of Agatston categories (Kappa and accuracy) and correlation of Agatston score (Spearman coefficient) between the AI model output and ground truth radiologists on non-gated CT.

| Subgroup | N | Kappa<br>(95% CI) | Accuracy<br>(95% CI) | Spearman $\rho$<br>(95% CI) |
| --- | --- | --- | --- | --- |
| **Sex** |  |  |  |  |
| Female | 272 | 0.957<br>(0.935, 0.975) | 91.5<br>(88.2, 94.8) | 0.965<br>(0.937, 0.985) |
| Male | 219 | 0.962<br>(0.936, 0.982) | 93.2<br>(89.8, 96.6) | 0.986<br>(0.974, 0.993) |
| **Age** |  |  |  |  |
| <65 years | 251 | 0.960<br>(0.938, 0.979) | 92.4<br>(89.1, 95.8) | 0.972<br>(0.949, 0.989) |
| >65 years | 240 | 0.958<br>(0.933, 0.978) | 92.1<br>(88.6, 95.5) | 0.973<br>(0.956, 0.987) |
| **Race** |  |  |  |  |
| Asian | 11 | 0.927<br>(0.818, 1.000) | 81.8<br>(58.7, 100.0) | 0.895<br>(0.561, 1.000) |
| Black or African American | 7 | 1.000<br>(1.000, 1.000) | 100.0<br>(100.0, 100.0) | 1.000<br>(1.000, 1.000) |
| White | 448 | 0.961<br>(0.943, 0.976) | 92.9<br>(90.5, 95.2) | 0.975<br>(0.961, 0.987) |
| Other | 11 | 0.855<br>(0.745, 0.964) | 63.6<br>(35.2, 92.9) | 0.930<br>(0.646, 1.000) |
| Declined | 12 | 1.000<br>(1.000, 1.000) | 100.0<br>(100.0, 100.0) | 0.993<br>(0.925, 1.000) |
| Unavailable | 2 | 1.000<br>(1.000, 1.000) | 100.0<br>(100.0, 100.0) | 1.000<br>(1.000, 1.000) |
| **Ethnicity** |  |  |  |  |
| Hispanic | 12 | 0.933<br>(0.833, 1.000) | 83.3<br>(62.0, 100.0) | 0.964<br>(0.826, 1.000) |
| Not Hispanic | 452 | 0.958<br>(0.941, 0.973) | 92.3<br>(89.9, 94.8) | 0.974<br>(0.960, 0.986) |
| Prefer not to say/Declined | 7 | 1.000<br>(1.000, 1.000) | 100.0<br>(100.0, 100.0) | 1.000<br>(1.000, 1.000) |
| Unavailable | 20 | 0.980<br>(0.940, 1.000) | 95.0<br>(85.4, 100.0) | 0.994<br>(0.959, 1.000) |
| **Manufacturer** |  |  |  |  |
| GE Medical Systems | 216 | 0.969<br>(0.944, 0.987) | 94.9<br>(91.9, 97.9) | 0.984<br>(0.969, 0.995) |
| Philips | 20 | 0.980<br>(0.940, 1.000) | 95.0<br>(85.2, 100.0) | 0.991<br>(0.958, 1.000) |
| Siemens | 214 | 0.948<br>(0.920, 0.970) | 89.7<br>(85.7, 93.9) | 0.962<br>(0.931, 0.983) |
| Toshiba | 41 | 0.961<br>(0.922, 0.990) | 90.2<br>(80.9, 99.1) | 0.979<br>(0.934, 0.998) |
| **Radiation dose** |  |  |  |  |
| Low dose | 269 | 0.967<br>(0.951, 0.982) | 92.9<br>(89.8, 96.0) | 0.977<br>(0.959, 0.990) |
| Routine dose | 222 | 0.950<br>(0.921, 0.973) | 91.4<br>(87.7, 95.1) | 0.972<br>(0.948, 0.989) |

