## Supplementary material for "Evaluation of an artificial intelligence model for opportunistic calculation of Agatston score on non-gated computed tomography of the chest": Figures (Supplementary)

**Figure S1:** Comparison of ground truth radiologists on non-gated CT with previously reported Agatston categories (A) and scores (B) from paired cardiac gated CT.

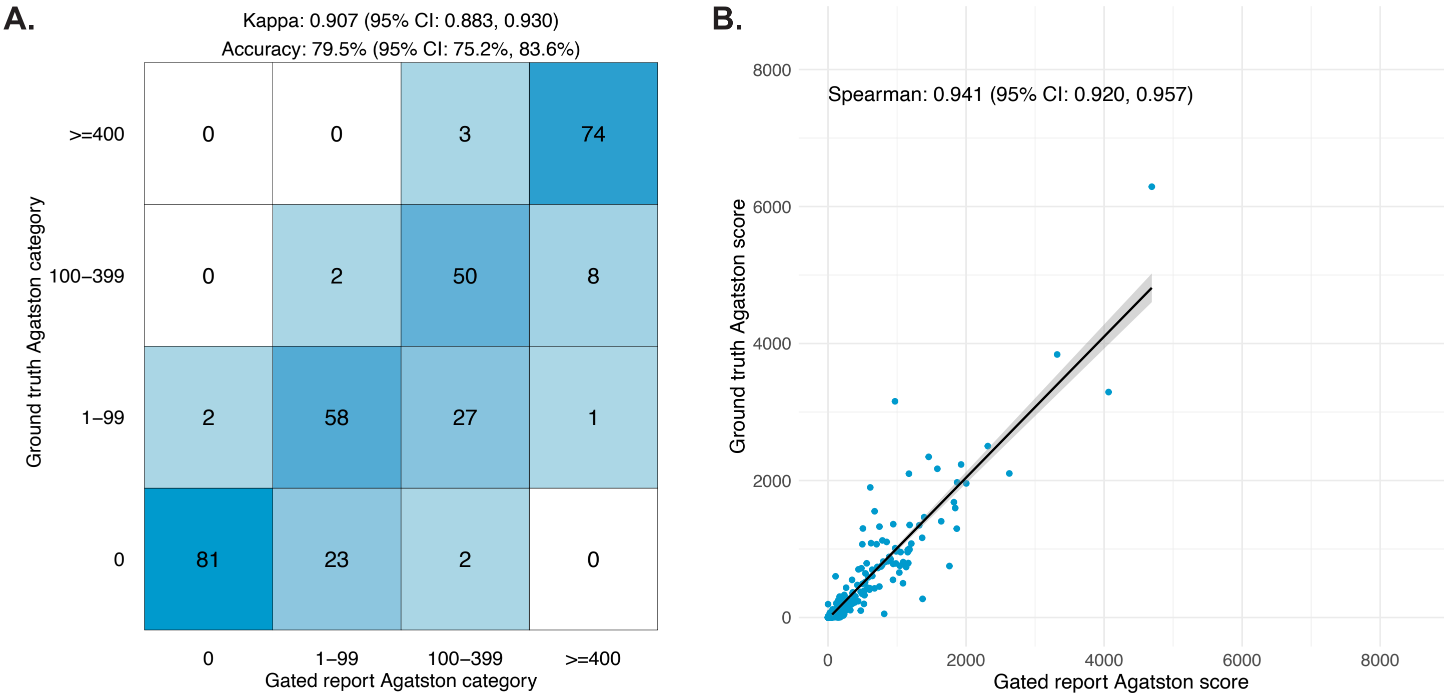
