## Supplementary material for "Evaluation of an artificial intelligence model for opportunistic calculation of Agatston score on non-gated computed tomography of the chest": Figures (Main text)

**Figure 1:** Cohort diagram.

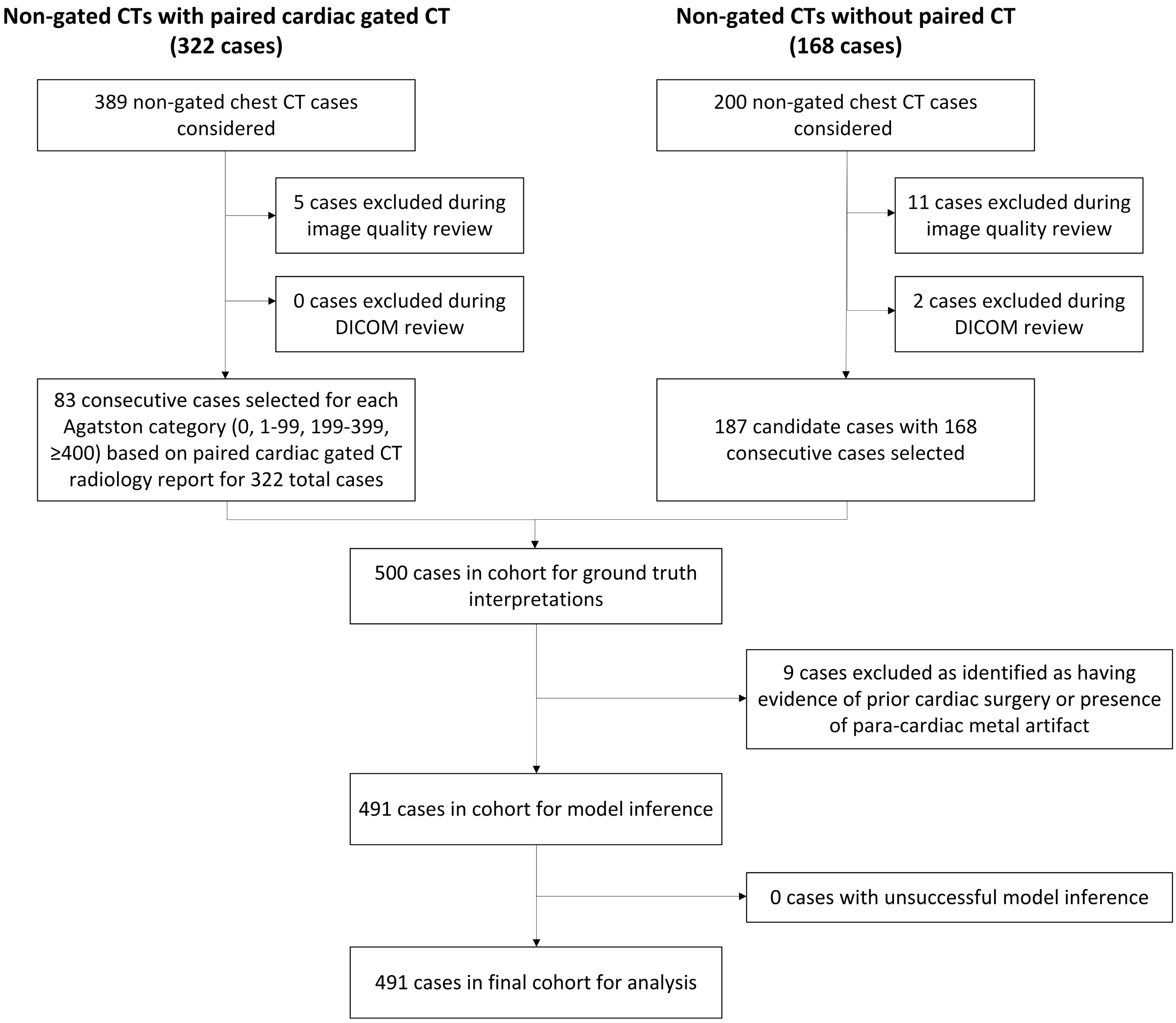

**Figure 2:** Comparison of AI model output compared with ground truth radiologists on non-gated CT for the Agatston category (A) and Agatston score (B).

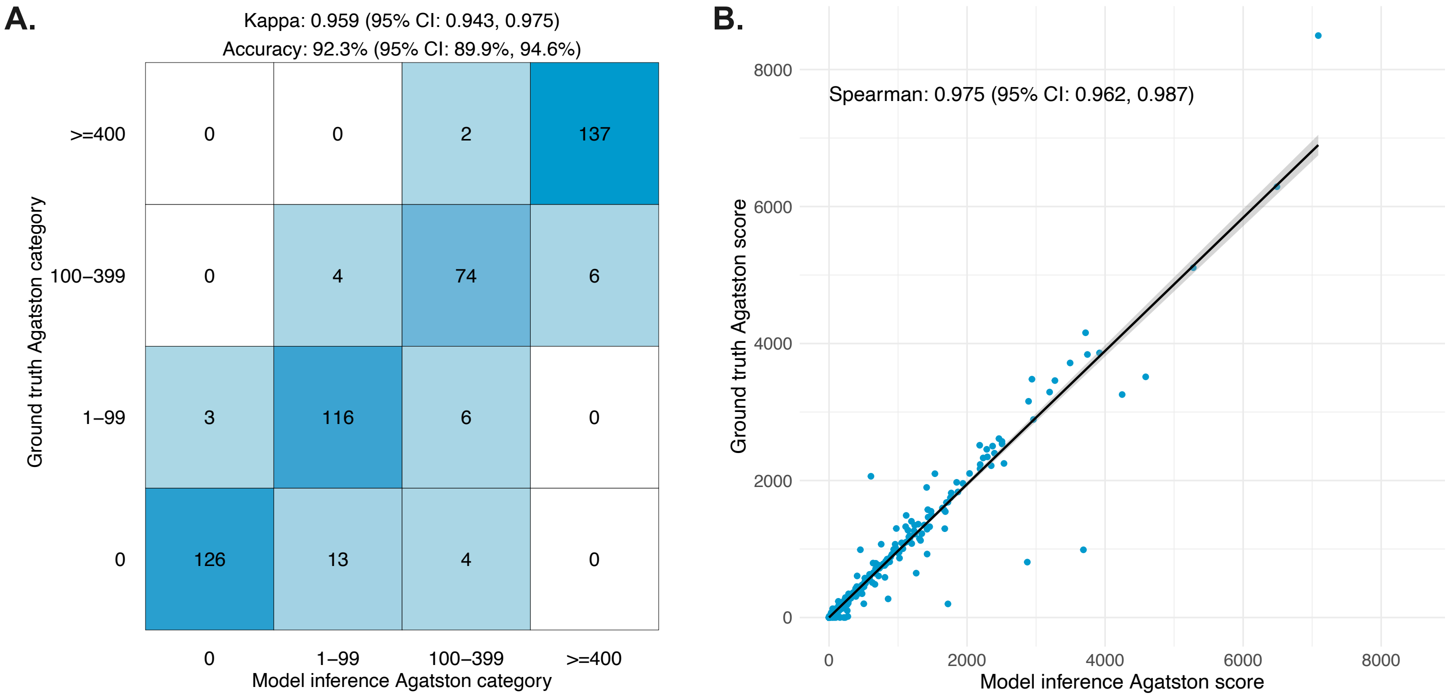

**Figure 3:** Scatterplots comparing AI model output with ground truth radiologists on non-gated CT for the Agatston score for each individual vessel including the right coronary artery (A), left anterior descending artery (B), left circumflex artery (C) and left main coronary artery (D).

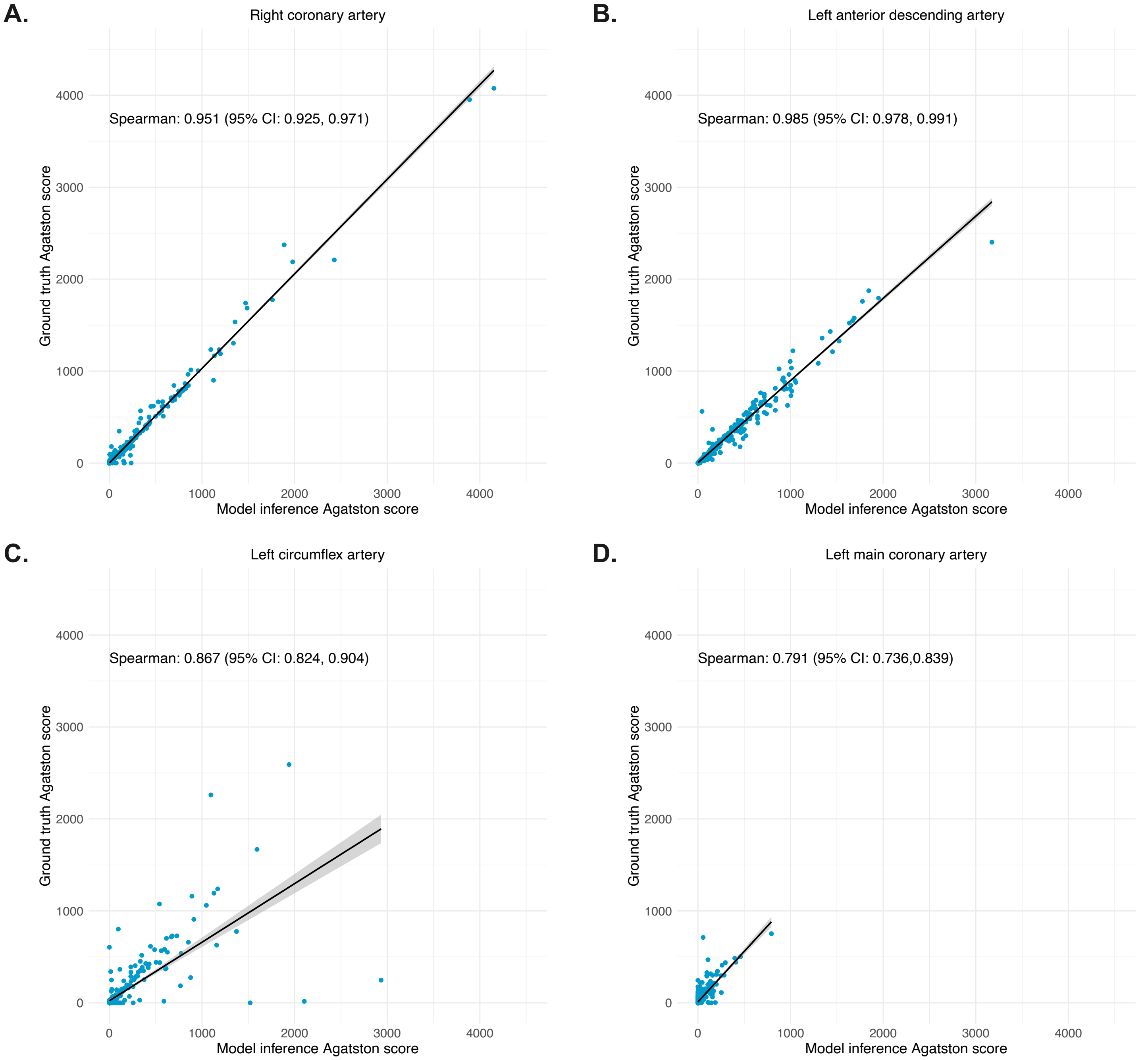

**Figure 4:** Comparison of AI model output on non-gated CT with previously reported Agatston categories (A) and scores (B) from paired cardiac gated CT.

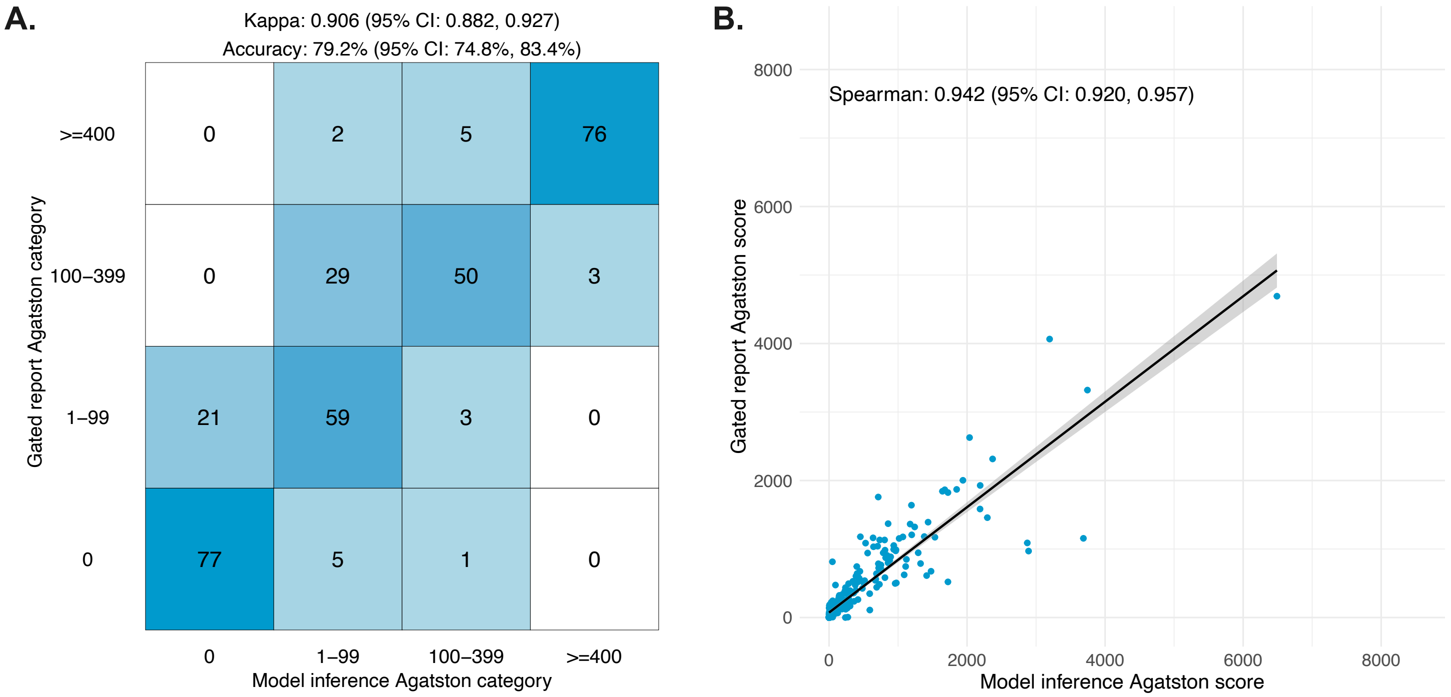

**Figure S1:** Comparison of ground truth radiologists on non-gated CT with previously reported Agatston categories (A) and scores (B) from paired cardiac gated CT.

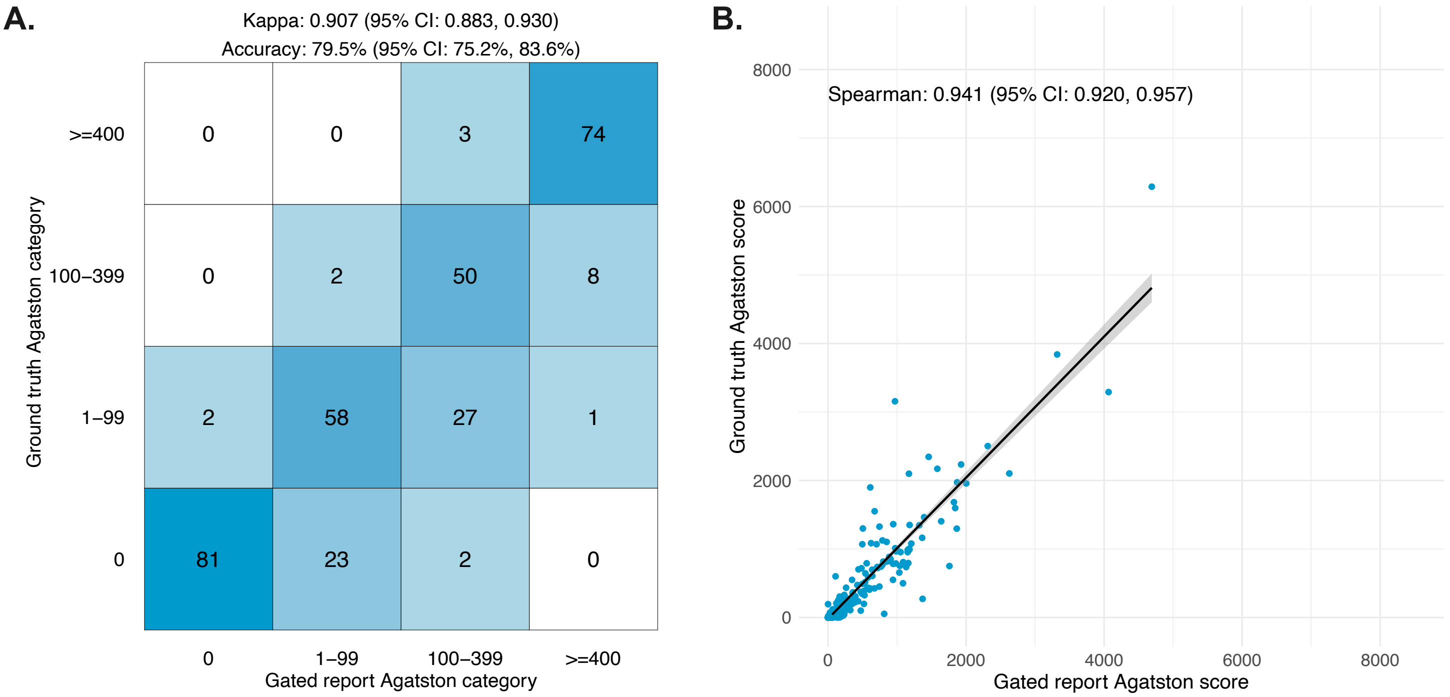
